# Trends and variations in Lithium usage across care settings in England between 2015-2024

**DOI:** 10.64898/2026.07.15.26357641

**Authors:** Hayley Schiffer, Louis Fisher, Helen J Curtis, Christopher Wood, Andrew Brown, Sebastian Bacon, Richard Croker, Ben Goldacre, Brian MacKenna, Victoria Speed, Orla Macdonald

**Author notes:** Corresponding authors: Orla Macdonald.

## Abstract

Lithium has been the gold standard for the treatment and prevention of relapse in bipolar disorder for over 60 years. Guidance from the National Institute for Health and Clinical Excellence states explicitly to “offer lithium as a first-line, long-term pharmacological treatment for bipolar disorder”. Yet, in the last two decades its use has been in decline with clinicians favouring anticonvulsants or antipsychotics when treating this condition.

In this study, we have used three openly available datasets containing prescribing data from primary and secondary care to explore trends in the use of lithium in England, showing both regional and temporal variance between 2015-2024.

We have shown that lithium use declined in primary care by 20.9% in the last ten years (2015-2024) and 10.9% overall in the last five years (2019 to 2025). We have also shown how there is some regional variation in the source of lithium for patients, although the vast majority is prescribed in primary care.

Further research into clinical behaviour is needed to understand what is driving the decrease in lithium usage, and what barriers and enablers may influence its use across the country.

## Introduction

Lithium salts have been used for psychiatric purposes since the mid 19th century and have been the treatment of choice for mood disorders since the 1950’s. Yet, since 2000, other antipsychotics and anticonvulsants have been licensed for mood disorders and gradually the relative rate of lithium use has declined.[1] This is despite evidence that lithium is superior for the prevention of relapse.[2][3] Since 2014, national guidelines have repeatedly stated that lithium should be offered as a first-line, long-term pharmacological treatment for bipolar disorder.[4][5]. Concerns about the underuse of lithium prompted the initiation of a national investigation, led by the Health Services Safety Investigations Body, in November 2025, to explore the use of lithium for the treatment of bipolar disorder, in England. [6]

Lithium is prescribed in both primary and secondary care settings. Open data describing the use of medicines in these settings within England exists in the form of: the English Prescribing Dataset (EPD)([7][8]; the *Hospital Prescribing Dispensed in the Community* dataset [9]; and the *Secondary Care Medicines* Data (SCMD) [10]. These data sources are described in **Box 1**. Combined, these three data sources allow for the analysis of national lithium prescribing across both primary and secondary care.

Using this data, we therefore set out to comprehensively describe trends in lithium usage across both primary and secondary care in England.

**Box 1.**
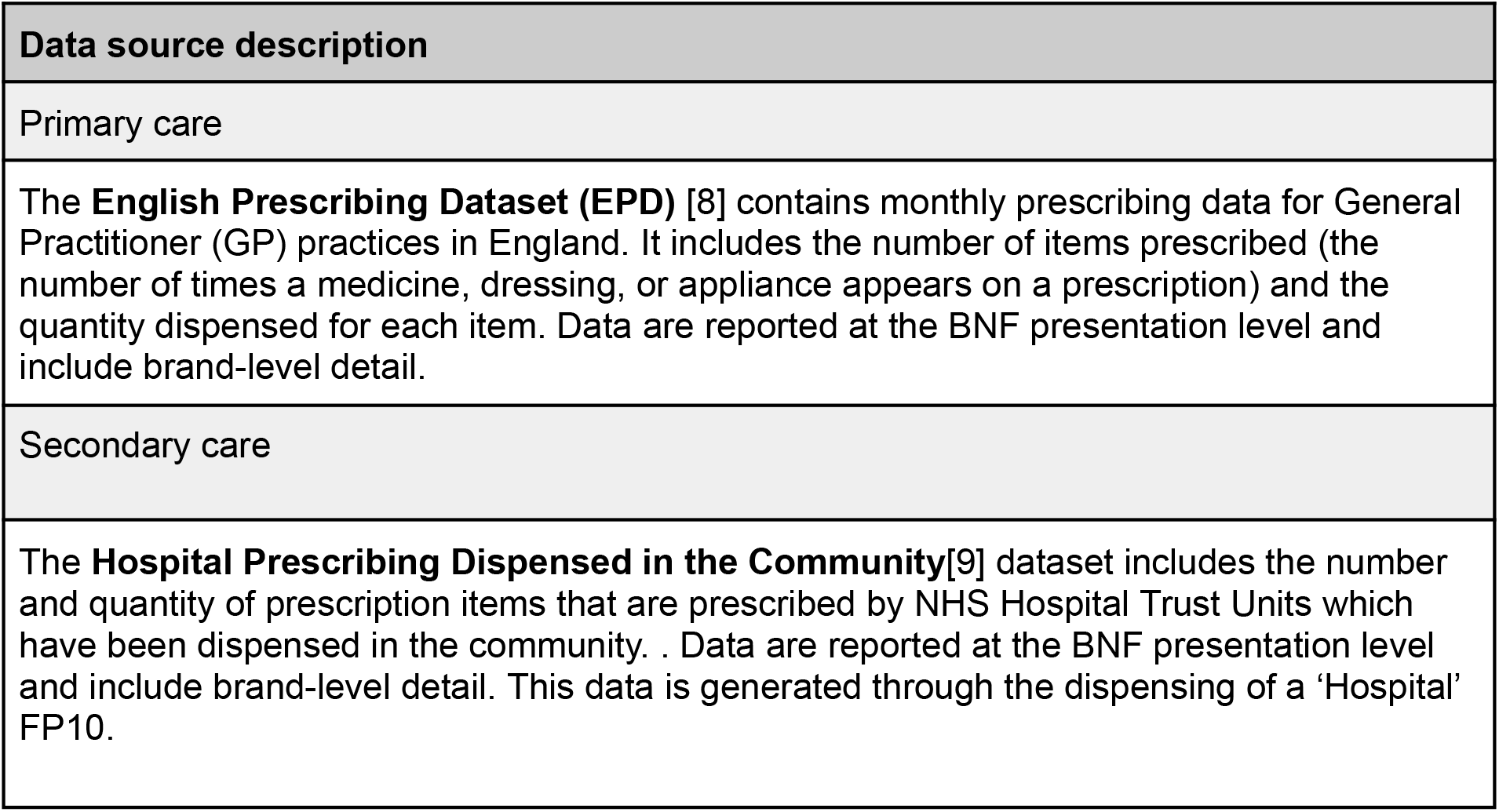
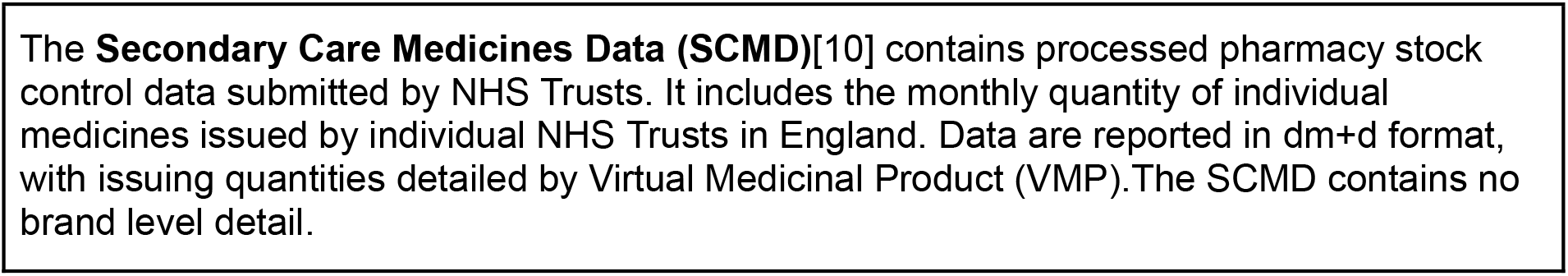
Description of openly available data sources for the analysis of medicines use in England. BNF: British National Formulary; dm+d: dictionary of medicines and devices.

## Methods

This study uses a retrospective observational design using the three openly available data sets outlined in Box 1. The EPD and *Hospital Prescribing Dispensed in the Community* dataset were accessed via the National Health Service Business Services Authority (NHSBSA) Open Data Portal.[11]. A processed version of the SCMD was accessed using the OpenPrescribing Hospitals platform, which supports rapid analyses of the SCMD.[12] The datasets contain data across different periods. We used the following time periods based on the availability of data: 2015-2024 for the EPD; 2017-2024 for the *Hospital Prescribing Dispensed in the Community*; and 2019-2024 for the SCMD.

In the EPD and Hospital Prescribing Dispensed in the Community datasets we identified all products using the BNF code prefix lithium carbonate (0402030K0) and lithium citrate (0402030P0). In the SCMD, we identified all products under the dm+d Virtual Therapeutic Moiety codes for lithium carbonate (776555000) and lithium citrate (776556004). Lithium chloride preparations were not included as this study focused on oral preparations only.

Within the described datasets, the quantity of lithium supplied is reported as a count of the number of tablets or volume of a liquid preparation. To allow aggregation of products of different strengths, the quantity of lithium reported in each data source was reported in Defined Daily Doses (DDDs). DDDs are an assumed average daily maintenance dose for drugs when used for their main indication in adults published by the World Health Organisation.[13] The DDD for lithium is defined as 24 mmol per day for the prophylaxis of mania or depression.[14] OpenPrescribing Hospitals supports the analysis of medicines usage in secondary care using DDDs natively so did not require any further calculation. To convert the quantity of lithium reported in the EPD and Hospitals Prescribing Dispensed in the Community datasets, the amount of lithium within each product reported was identified using the dictionary of medicines and devices (dm+d)[15] (**Table S1**) and used to calculate the quantity of lithium in grams. This was converted to the quantity in mmol using the equivalence described within the Summary of Product Characteristics for a sample product (lithium citrate - 94.26mg:1mmol; lithium carbonate - 37.04mg:1mmol).[16] This was finally converted to the quantity in DDDs by dividing the quantity in mmols by the DDD for lithium.[17]

To investigate regional variation in lithium prescribing, we calculated lithium use per capita for each NHS region. We used annual population estimates for each region from the Office for National Statistics (ONS).[18] Regional trends in lithium usage per capita, within each care setting, were plotted between 2019–2024.

Data management was performed using Python with analysis carried out using R. All code for data management and analysis is archived online at https://github.com/bennettoxford/lithium_project25.

## Results

We identified 18 lithium preparations within the primary care and community prescribing datasets and 7 Virtual Medicinal Products within the secondary care data (**Table S2**). **Table 1** shows the total number of DDDs of lithium used, within the respective earliest and latest year for each data source.

**Table 1.**
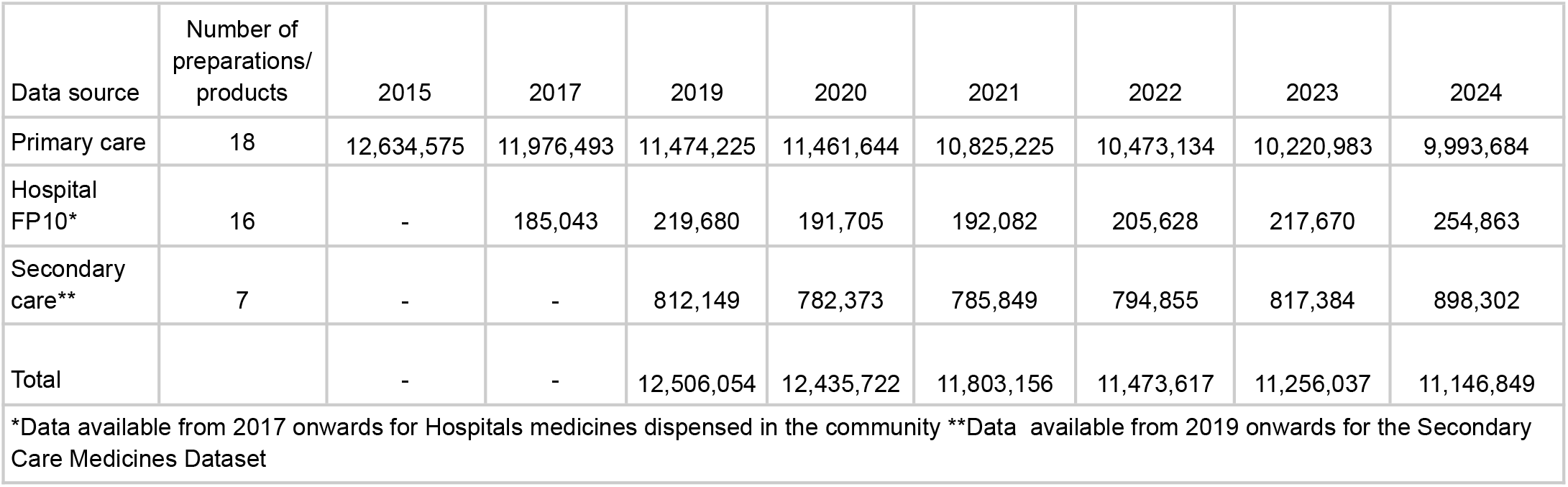
Summary of the number of lithium products identified in each source and the total number of DDDs reported in each source between 2015 and 2024.

The majority of lithium is prescribed in primary care. In 2024, 9,993,684 (89.7%) DDDs were dispensed from primary care whereas only 898,302 (8.1%) DDDs were issued in hospitals and 254,863 (2.3%%) DDDs dispensed from hospital FP10s. Further details are shown in **Table S3**.

Total lithium usage across settings between 2015 and 2024 is shown in **Figure 1**. This shows a steady decline in the use of lithium over this period. Usage in primary care decreased from 12,634,575 to 9,993,684 (-20.9%) between 2015 and 2024. This is contrasted by trends seen in secondary care which represent a much smaller proportion of total lithium use. Prescribing on FP10s in secondary care has increased from 185,043 to 254,863 (+37.7%) between 2017 and 2024, and use within hospitals has increased by 10.6%, from 812,149 in 2019 to 898,302 in 2024. Despite increases in secondary care use, the total number of DDDs being used each year has decreased from 12,506,054 to 11,146,849 (-10.9%) between 2019 and 2024.

**Figure 1.**
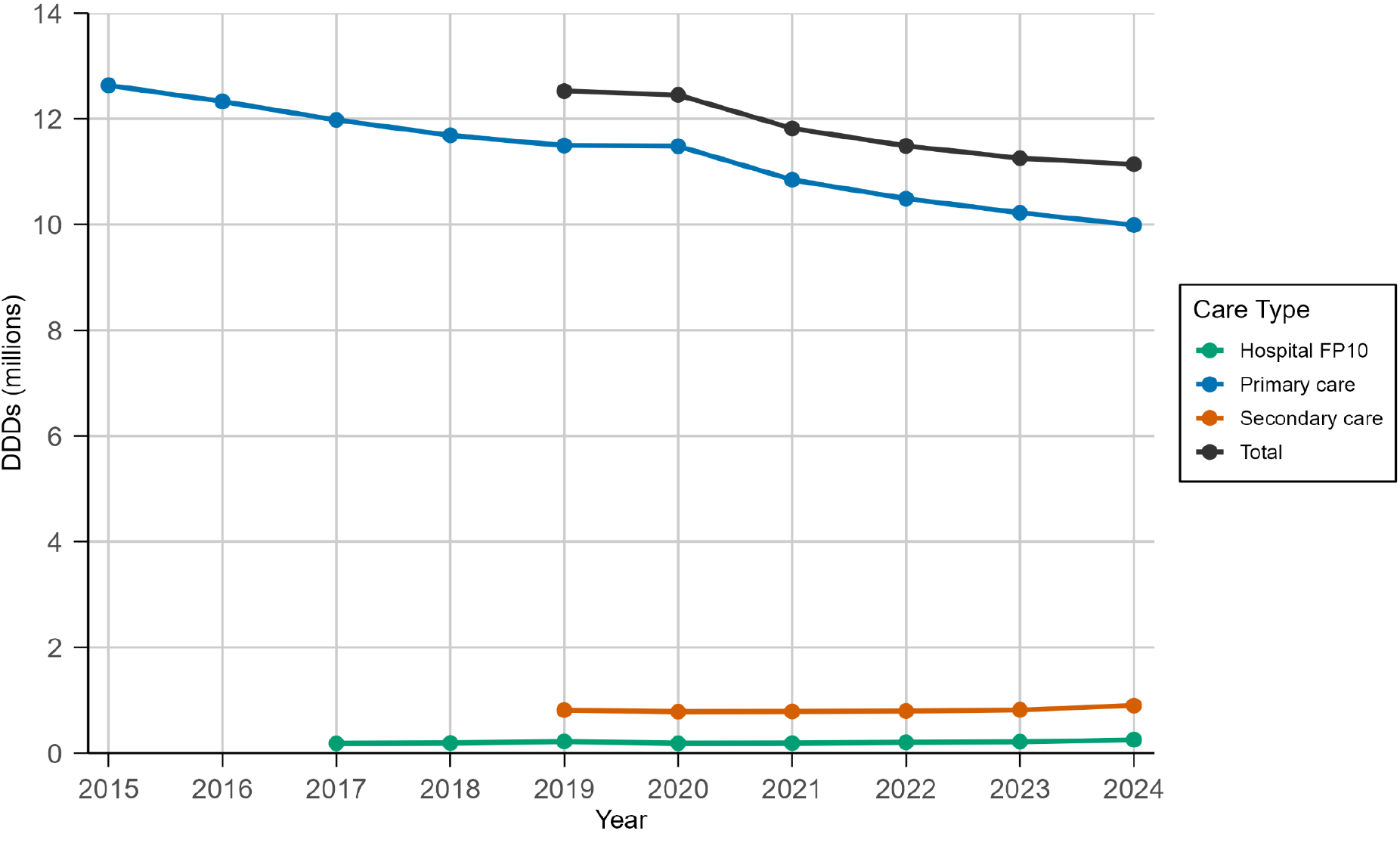
Total number of DDDs (millions) of lithium used by source, in England, between 2015-2024.

Over the observed 5-year period from 2019 to 2024, most regions saw a gradual decrease in lithium use per capita, apart from London which showed a small increase in lithium use from 189.5 in 2023 to 193.2 DDDs/1000 in 2024 (**Figure 2, Table S4**). The North-West has seen the greatest decrease in lithium use the past 5 years, decreasing from 260.74 in 2019 to 207.86 DDDs/1000 in 2024 (-20.3%).

**Figure 2.**
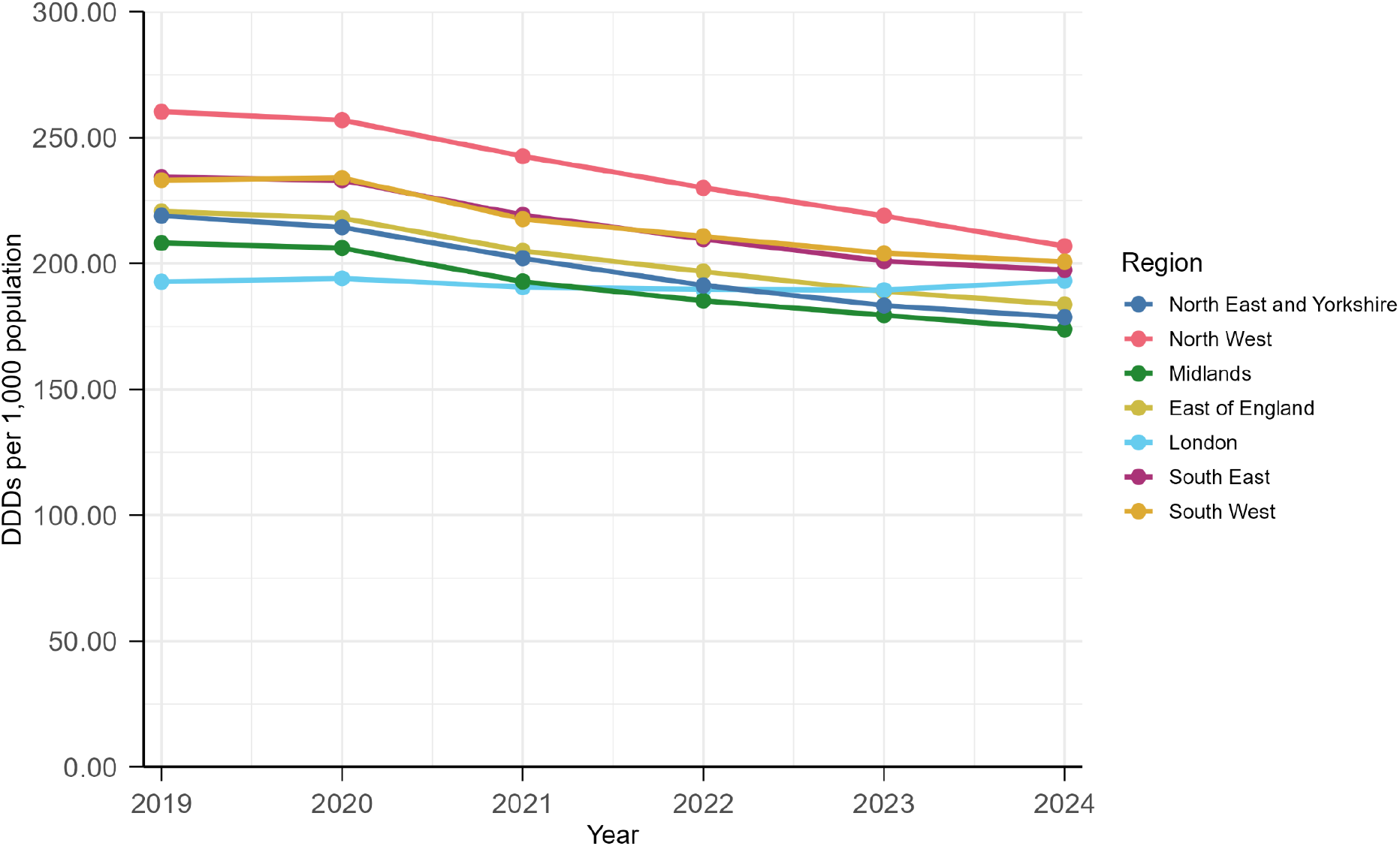
DDDs of lithium per 1000 population reported across all sources between 2019-2024 within each NHS region in England.

We did not find marked regional variation in the total quantity of lithium used across England, although there was some notable variation in the prescribing source of lithium. **Figure 3** shows the total DDDs per 1000 of lithium supplied from each source, in 2024. Annual rates of lithium use for each data source are shown in **Table S5-S7**.

**Figure 3.**
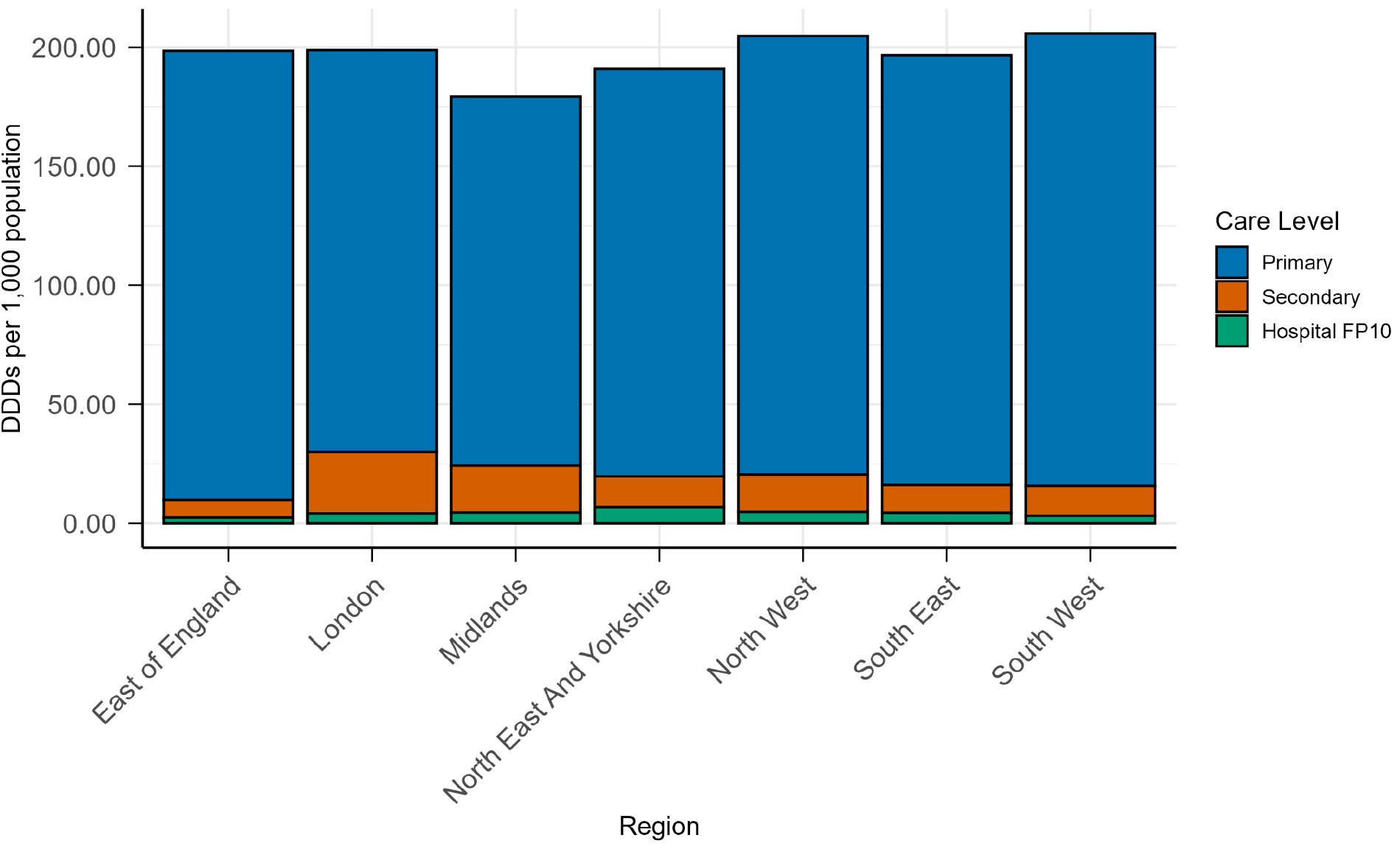
DDDs of lithium per 1000 population reported within each NHS region in each data source in 2024

**Figure 4.**
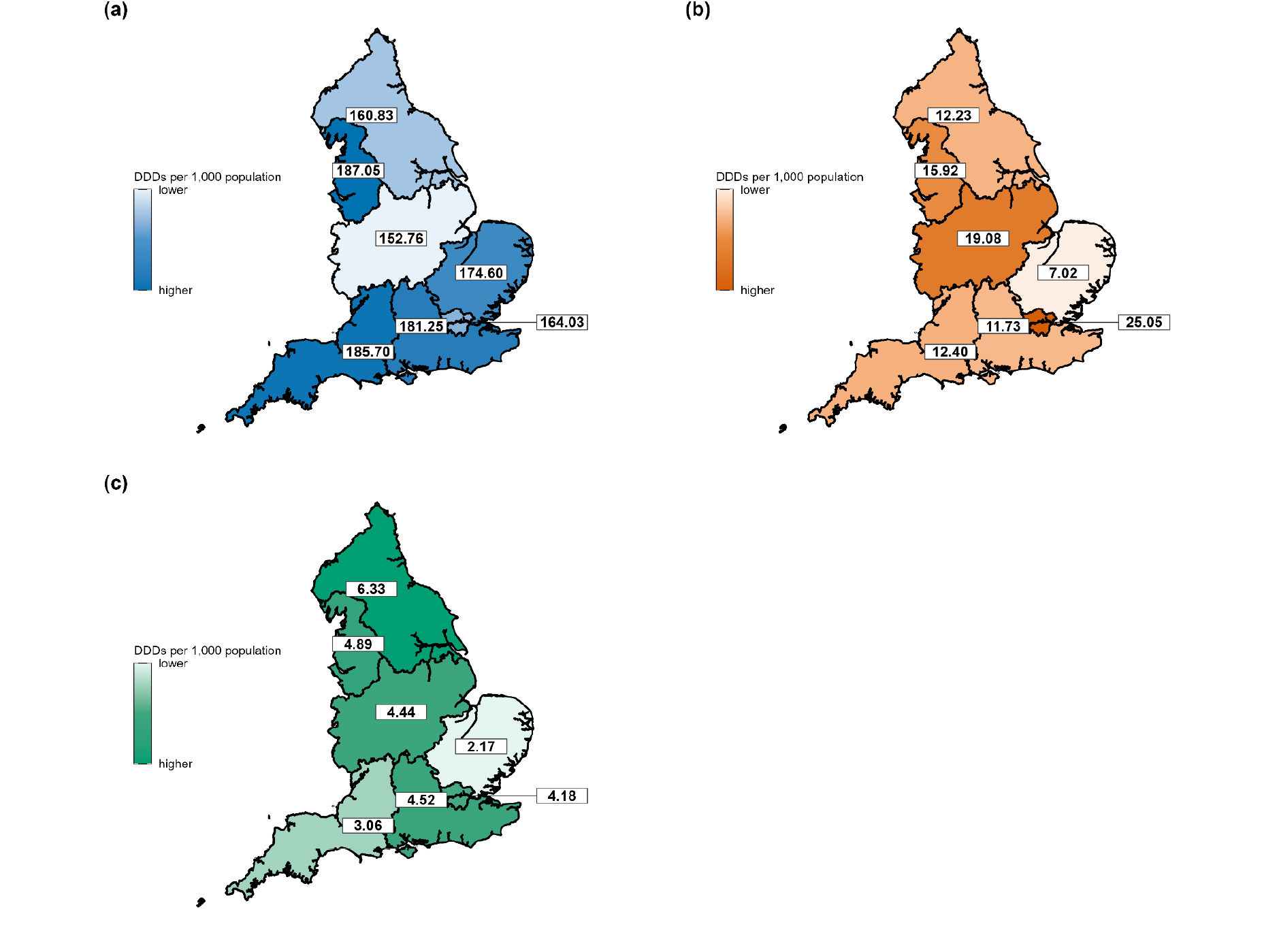
DDDs of lithium per 1000 population reported in 2024 in a) primary care, b) secondary care, c) hospital FP10.

In primary care, the highest DDD per 1000 was observed in the North West (187.05), followed by the South-West (185.70) and the lowest prescribing rates in primary care was recorded in the Midlands (152.76). In secondary care, the highest DDD per 1000 was in London (25.05), followed by the Midlands (19.08), while the East of England had the lowest secondary care amount issued (7.02). Where a hospital FP10 was the source of the lithium, the highest DDDs per 1000 were in the North East and Yorkshire (6.33), followed by the North West (4.89) and the lowest was in the East of England (2.17).

## Discussion

Our study has shown that lithium usage has continued to decline in England over the last 10 years, despite national guidelines recommending that it is used first line for the prevention of relapse in bipolar disorder.[5,19] Furthermore, we have shown that there is some regional variation in the source of lithium for patients. The majority of lithium is prescribed and dispensed in primary care, but within some areas, a greater proportion of lithium prescriptions comes from secondary care providers. Supply of lithium from hospital sources appears to have increased within the last five years.This shift is in contrast with the recently published NHS 10 year plan, which strongly supports moving care from hospital to the community for people with mental illness.[20]

### Strengths and weaknesses of the study

To our knowledge, this is the first study to combine the openly available EPD, SCMD and Hospital Prescribing Dispensed in the Community datasets to explore complete prescribing trends of a medicine within the NHS in England. As a result, we have been able to comprehensively describe the regional and temporal variations of lithium use in the country. However, there are limitations to this data. None of the datasets included within this analysis include patient level data, therefore we cannot describe how many patients were treated with lithium within each region, nor identify the severity or details of their illness. Additionally, the SCMD reflects processed pharmacy stock rather than individual patient use. As such, we are not able to differentiate between lithium that is issued but not administered to that which is given to a patient within secondary care. Finally, due to the absence of patient level data in these datasets, we have not been able to explore factors that are driving this persistent decrease in usage.

### Findings in context

Bipolar disorder is a severe, chronic mental illness affecting between 1-2% of the population in the UK.[21] Multiple studies have shown that lithium is the most effective treatment for preventing relapse in this disorder [2,3] and that population level outcomes of bipolar disorder could be improved by increasing the number of patients who are taking lithium.[22]

In 2021, Ng et al. explored the prevalence of bipolar disorder and psychotropic medication utilisation within a small subset of UK patients from primary care only from 2001 to 2018 and reported a 14.58% decline in the use of lithium in the UK over that period, with a concurrent increase in antipsychotic prescribing.[23] Our study, across the entire population, has shown that the majority of lithium is dispensed from primary care prescriptions, and there is a continuing pattern of decline in its use.

In 2023, a national audit of lithium monitoring was conducted across the UK. 63 secondary care trusts, representing the majority of mental health trusts in England, submitted data on patients prescribed lithium. [24] The *Priadel* brand of lithium carbonate was prescribed in 84% of cases and either bipolar disorder or affective disorders were diagnosed in 79% of patients. Despite the breadth of data collected by this audit, it did not report on regional variations in lithium provision.

### Future research and implications for practice

There are unanswered questions about why lithium use continues to decline, despite robust evidence confirming its superiority in reducing relapse in bipolar disorder.[2,3,5,22] Researchers and commissioners might want to consider what systems promote lithium use. Do lithium databases increase confidence in safe outcomes, thus promoting prescribing? Is it more advantageous to manage lithium within secondary care, for example through lithium clinics, or does primary care prescribing improve access and thereby increase patient willingness to use the drug?

Our group has developed two openly available tools to support easier analysis of the data sources used in this study: OpenPrescribing and OpenPrescribing Hospitals.[12,25] Both platforms enable users to explore how lithium is prescribed in their region and to measure the impact of changes in service or quality improvement projects.

Lithium is an old drug and yet one that has a superior effect on patient outcomes in bipolar disorder. We need to ensure that our healthcare systems promote its use where possible, to give our patients the best chance of recovery.

## Data Availability

All data produced, along with the code for data management and analysis, are available online at https://github.com/bennettoxford/lithium_project25.

https://docs.google.com/document/d/1eNEXTxeC9NHoC1yv2WDYpc7JCyq7xfMPssa9e92eR5c/edit?tab=t.0

## Ethical approval

This study uses exclusively open, publicly available data, therefore no ethical approval was required.

## Guarantor

VS is guarantor.

## Contributorship

**Conceptualization**: Hayley Schiffer, Orla Macdonald, Brian MacKenna, Victoria Speed

**Data curation**: Louis Fisher, Richard Croker, Christoper Wood, Hayler Schiffer

**Formal analysis**: Hayley Schiffer, Louis Fisher

**Funding acquisition**: Ben Goldacre and Brian MacKenna.

**Investigation**: Hayley Schiffer, Orla Macdonald, Victoria Speed.

**Methodology**: Hayley Schiffer, Orla Macdonald, Victoria Speed, Louis Fisher, Richard Croker, Ben Goldacre, and Brian MacKenna.

**Project administration**: Victoria Speed

**Resources**: Sebastian Bacon.

**Software**: Louis Fisher and Sebastian Bacon.

**Supervision**: Ben Goldacre.

**Validation**: Hayley Schiffer, Orla Macdonald, Louis Fisher, Christopher Wood, Richard Croker, and Victoria Speed.

**Visualization**: Hayley Schiffer, Orla Macdonald, Louis Fisher, Ben Goldacre, and Victoria Speed.

**Writing - original draft**: Hayley Schiffer, Orla Macdonald, Louis Fisher, Victoria Speed.

**Writing - review & editing**: Hayley Schiffer, Orla Macdonald, Louis Fisher, Helen J. Curtis, Christopher Wood, Sebastian Bacon, Richard Croker, Ben Goldacre, Brian MacKenna, and Victoria Speed.

