## Supplementary data for "Trends and variations in Lithium usage across care settings in England between 2015-2024"

**Table S1.** Product information for products identified in the English Prescribing Dataset (EPD) and the Hospitals Prescribing Dispensed in the Community dataset, identified using the dictionary of medicines and devices (dm+d).

| BNF Code | BNF Name | Chemical | Strength numerator (mg) | Strength denominator (ml) |
|---|---|---|---|---|
| 0402030K0AAACAC | Lithium carbonate 250mg tablets | Lithium Carbonate | 250 | |
| 0402030K0AAAFAF | Lithium carbonate 400mg modified-release tablets | Lithium Carbonate | 400 | |
| 0402030K0AAAGAG | Lithium carbonate 450mg modified-release tablets | Lithium Carbonate | 450 | |
| 0402030K0AAAIAI | Lithium carbonate 200mg modified-release tablets | Lithium Carbonate | 200 | |
| 0402030K0AAAPAP | Lithium carbonate 200mg/5ml oral suspension | Lithium Carbonate | 40 | 1 |
| 0402030K0BBAAAC | Camcolit 250 tablets | Lithium Carbonate | 250 | |
| 0402030K0BBABAF | Camcolit 400 modified-release tablets | Lithium Carbonate | 400 | |
| 0402030K0BDAAAG | Liskonum 450mg modified-release tablets | Lithium Carbonate | 450 | |
| 0402030K0BFAAAF | Priadel 400mg modified-release tablets | Lithium Carbonate | 400 | |
| 0402030K0BFABAI | Priadel 200mg modified-release tablets | Lithium Carbonate | 200 | |
| 0402030K0BGAAAF | Lithonate 400mg modified-release tablets | Lithium Carbonate | 400 | |

| | | | | |
|---|---|---|---|---|
| 0402030P0AAAIAI | Lithium citrate 520mg/5ml oral solution sugar free | Lithium Citrate | 104 | 1 |
| 0402030P0AAAJAJ | Lithium citrate 10.8mmol/5ml oral solution sugar free | Lithium Citrate | 203.6 | 1 |
| 0402030P0AAAKAK | Lithium citrate 1.018g/5ml oral solution | Lithium Citrate | 203.6 | 1 |
| 0402030P0AAALAL | Lithium citrate 509mg/5ml oral solution | Lithium Citrate | 101.8 | 1 |
| 0402030P0BCAAAI | Priadel 520mg/5ml liquid | Lithium Citrate | 104 | 1 |
| 0402030P0BDAAAL | Li-Liquid 509mg/5ml oral solution | Lithium Citrate | 101.8 | 1 |
| 0402030P0BDABAK | Li-Liquid 1.018g/5ml oral solution | Lithium Citrate | 203.6 | 1 |

**Table S2.** Products identified within each data source and the total number of DDDs reported by each data source in their respective earliest and latest date.

| Primary care/community prescribing (FP10) | | | | |
|---|---|---|---|---|
| | Total DDDs | | | |
| | Primary care | | Community prescribing (FP10) | |
| BNF name (BNF code) | 2015 | 2024 | 2017 | 2024 |
| Total (all products) | 12,634,575 | 9,993,684 | 185,043 | 254,863 |
| Priadel 400mg modified-release tablets (0402030K0BFAAAF) | 7,432,995 | 6,196,653 | 81,233 | 133,176 |
| Priadel 200mg modified-release tablets (0402030K0BFABAI) | 2,434,868 | 2,524,442 | 32,297 | 47,514 |
| Lithium carbonate 400mg modified-release tablets (0402030K0AAAFAF) | 1,367,066 | 430,551 | 47,520 | 47,702 |
| Camcolit 400 modified-release tablets (0402030K0BBABAF) | 302,127 | 321,520 | 975 | 643 |
| Lithium carbonate 200mg modified-release tablets (0402030K0AAAIAI) | 447,674 | 169,317 | 17,404 | 17,011 |
| Lithium carbonate 250mg tablets (0402030K0AAACAC) | 102,800 | 144,428 | 2,250 | 3,046 |
| Priadel 520mg/5ml liquid | 89,971 | 81,617 | 1,116 | 2,146 |

| | | | | |
|---|---|---|---|---|
| (0402030P0BCAAAI) | | | | |
| Liskonum 450mg modified-release tablets (0402030K0BDAAAG) | 102,223 | 67,306 | 177 | 1,095 |
| Lithium carbonate 450mg modified-release tablets (0402030K0AAAGAG) | 43,945 | 21,898 | 209 | 847 |
| Camcolit 250 tablets (0402030K0BBAAAC) | 248,178 | 0 | 317 | 8 |
| Lithium citrate 520mg/5ml oral solution sugar free (0402030P0AAAIAI) | 28,522 | 9,573 | 666 | 842 |
| Li-Liquid 509mg/5ml oral solution (0402030P0BDAAAL) | 10,620 | 14,181 | 93 | 118 |
| Lithium citrate 509mg/5ml oral solution (0402030P0AAALAL) | 10,880 | 4,406 | 319 | 479 |
| Li-Liquid 1.018g/5ml oral solution (0402030P0BDABAK) | 4,719 | 6,062 | 131 | 44 |
| Lithium citrate 1.018g/5ml oral solution (0402030P0AAAKAK) | 5,748 | 1,644 | 243 | 34 |
| Lithium carbonate 200mg/5ml oral suspension (0402030K0AAAPAP) | 292 | 77 | 92 | 157 |
| Lithonate 400mg modified-release tablets (0402030K0BGAAAF) | 1,925 | 9 | 0 | 0 |
| Lithium citrate 10.8mmol/5ml oral solution sugar free (0402030P0AAAJAJ) | 22 | 0 | 0 | 0 |

| Secondary care | | |
|---|---|---|
| VMP Name | Total DDDs | |
| | 2019 | 2024 |
| Total (all products) | 812,149 | 898,302 |
| Lithium carbonate 400mg modified-release tablets (39110111000001104) | 571,839 | 628,061 |
| Lithium carbonate 200mg modified-release tablets (39110011000001104) | 182,834 | 207,769 |
| Lithium citrate 520mg/5ml oral solution sugar free (36040311000001104) | 39,928 | 50,368 |
| Lithium citrate 509mg/5ml oral solution (42295911000001104) | 8,286 | 3,840 |
| Lithium carbonate 250mg tablets (42295711000001104) | 3,754 | 3,831 |

**Table S3** Summary of the number of lithium products identified in each source and the total number of DDDs reported in each source between 2015 and 2024. Full detail

| Data source | Number of preparation s/products | 2015 | 2016 | 2017 | 2018 | 2019 | 2020 | 2021 | 2022 | 2023 | 2024 |
|---|---|---|---|---|---|---|---|---|---|---|---|
| Primary care | 18 | 12,634,575 | 12,332,939 | 11,976,493 | 11,669,2553 | 11,474,225 | 11,461,644 | 10,825,225 | 10,473,134 | 10,220,983 | 9,993,684 |
| Hospital FP10* | 16 | - | - | 185,043 | 190,609 | 219,680 | 191,705 | 192,082 | 205,628 | 217,670 | 254,863 |
| Secondary care** | 7 | - | - | - | - | 812,149 | 782,373 | 785,849 | 794,855 | 817,384 | 898,302 |
| Total | | - | - | - | - | 12,506,054 | 12,435,722 | 11,803,156 | 11,473,617 | 11,256,037 | 11,146,849 |
| *Data available from 2017 onwards for Hospitals medicines dispensed in the community **Data available from 2019 onwards for the Secondary Care Medicines Dataset | | | | | | | | | | | |

**Table S4:** DDDs of lithium per 1000 patients within each NHS region in England across all settings between 2019-2024.

| Region | DDDs of lithium per 1000 patients | | | | | |
|---|---|---|---|---|---|---|
| | 2019 | 2020 | 2021 | 2022 | 2023 | 2024 |
| North East and Yorkshire | 219.43 | 215.17 | 202.8 | 191.86 | 183.88 | 179.39 |
| North West | 260.74 | 257.36 | 243.03 | 230.48 | 219.55 | 207.86 |
| Midlands | 209.33 | 206.96 | 193.54 | 186.23 | 181.56 | 176.28 |
| East of England | 220.75 | 218.35 | 205.06 | 196.87 | 189.03 | 183.79 |
| London | 192.78 | 194.24 | 190.75 | 189.87 | 189.57 | 193.26 |
| South East | 234.16 | 232.96 | 219.36 | 209.79 | 201.04 | 197.5 |
| South West | 233.45 | 234.52 | 218.02 | 210.95 | 204.46 | 201.16 |

**Table S5:** DDDs of lithium per 1000 patients within each NHS region in England in primary care between 2015-2024.

| Region | DDDs of lithium per 1000 patients | | | | | | | | | |
|---|---|---|---|---|---|---|---|---|---|---|
| | 2015 | 2016 | 2017 | 2018 | 2019 | 2020 | 2021 | 2022 | 2023 | 2024 |
| North East and Yorkshire | 235.39 | 226.38 | 217.27 | 207.42 | 200.73 | 198.58 | 185.79 | 176.5 | 167.85 | 160.83 |
| North West | 273.13 | 267.13 | 258.6 | 249.9 | 241.51 | 239.04 | 223.76 | 210.92 | 200.19 | 187.05 |
| Midlands | 212.29 | 204.75 | 196.7 | 190.92 | 187.39 | 185.99 | 173.18 | 164.96 | 159.35 | 152.76 |
| East of England | 240.43 | 230.28 | 222.49 | 216.47 | 211.14 | 210.02 | 197.35 | 188.28 | 180.26 | 174.6 |
| London | 187.49 | 180.18 | 173.06 | 168.84 | 169.33 | 170.65 | 165.81 | 163.99 | 162.71 | 164.03 |
| South East | 234.46 | 229.06 | 223.2 | 221.63 | 217.65 | 217.63 | 204.92 | 195.5 | 187.26 | 181.25 |
| South West | 248.09 | 240.64 | 234.33 | 223.91 | 218.57 | 220.8 | 204.76 | 197.48 | 190.85 | 185.7 |

**Table S6:** DDDs of lithium per 1000 patients within each NHS region in England in secondary care between 2019-2024.

| Region | DDDs of lithium per 1000 patients | | | | | |
|---|---|---|---|---|---|---|
| | 2019 | 2020 | 2021 | 2022 | 2023 | 2024 |
| North East and Yorkshire | 13.95 | 12.09 | 12.2 | 10.69 | 10.87 | 12.23 |
| North West | 15.16 | 15.13 | 15.41 | 15.45 | 15.22 | 15.92 |
| Midlands | 16.64 | 16.47 | 16.2 | 16.91 | 17.81 | 19.08 |
| East of England | 7.79 | 6.91 | 6.45 | 6.89 | 6.9 | 7.02 |
| London | 19.95 | 20.04 | 21.39 | 22.13 | 22.86 | 25.05 |
| South East | 12.76 | 12.07 | 11.23 | 10.75 | 10.25 | 11.73 |
| South West | 11.8 | 11.47 | 11.43 | 11.53 | 11.47 | 12.4 |

**Table S7.** DDDs of lithium per 1000 patients within each NHS region in England in secondary care FP10 data between 2017-2024.

| Region | DDDs of lithium per 1000 patients | | | | | | | |
|---|---|---|---|---|---|---|---|---|
| | 2017 | 2018 | 2019 | 2020 | 2021 | 2022 | 2023 | 2024 |
| North East and Yorkshire | 4.28 | 4.07 | 4.75 | 4.5 | 4.81 | 4.67 | 5.16 | 6.33 |
| North West | 2.93 | 3.27 | 4.07 | 3.19 | 3.86 | 4.11 | 4.14 | 4.89 |
| Midlands | 4.27 | 4.79 | 5.3 | 4.5 | 4.16 | 4.36 | 4.4 | 4.44 |
| East of England | 1.67 | 1.69 | 1.82 | 1.42 | 1.26 | 1.7 | 1.87 | 2.17 |
| London | 3.05 | 3.13 | 3.5 | 3.55 | 3.55 | 3.75 | 4 | 4.18 |
| South East | 3.23 | 3.35 | 3.75 | 3.26 | 3.21 | 3.54 | 3.53 | 4.52 |
| South West | 3.02 | 2.44 | 3.08 | 2.25 | 1.83 | 1.94 | 2.14 | 3.06 |
